## Supplement for "Implementing evidence ecosystems in the public health service: Development of a seven-step framework for designing tailored training programs"

*Table S1. Search strategy for the database PUBMED*

| |
|---|
| ((((Public Health Administration[MeSH Terms] OR Public Health Practice[MeSH Terms] OR Personnel Turnover[MeSH Terms] OR Public Health Systems Research[MeSH Terms] OR Schools, Public Health[MeSH Terms]) AND (train*[Title] OR educat*[Title] OR postgrad*[Title] OR post-grad*[Title] OR specialt*[Title]))) OR ((public health[Title] OR health authori*[Title] OR health department*[Title]) AND ((Health Workforce[MeSH Terms] OR Capacity Building[MeSH Terms] OR Personnel Management[MeSH Terms] OR Education, Public Health Professional[MeSH Terms]) OR (train*[Title] OR educat*[Title] OR postgrad*[Title] OR post-grad*[Title] OR specialt*[Title] OR (capacity[Title] AND building[Title]) OR specialty[Title] OR workforce*[Title] OR manpower[Title] OR competenc*[Title] OR abilit*[Title] OR skill*[Title] OR curricul*[Title] OR learning goals[Title] OR residenc*[Title]))) OR (((public health[Title] OR health authori*[Title] OR health department*[Title]) OR (public health workforce[Title/Abstract] OR epidemiolog*[Title/Abstract] OR Public health special*[Title/Abstract])) AND (capacity building[Title/Abstract] OR capacity building[MeSH Terms] OR (capacity[Title/Abstract] AND building[Title/Abstract]) OR specialt*[Title] OR postgrad*[Title/Abstract] OR post-grad*[Title/Abstract] OR program*[Title/Abstract]) AND (educat*[Title/Abstract] OR train*[Title/Abstract]))) AND (((Switzerland* OR Swiss* OR Schweiz*) OR (Aargau OR Appenzell Outer-Rhodes OR Appenzell Ausser-Rhoden OR Appenzell Inner-Rhodes OR Appenzell Inner-Rhoden OR Basel OR Basel District OR Basel Landschaft OR Bern OR Bern OR Fribourg OR Freiburg OR Geneva OR Genf OR Glarus OR Glarus OR Grisons OR Graubünden OR Jura OR Jura OR Lucerne OR Luzern OR Neuchâtel OR Neuenburg OR Nidwalden OR Obwalden OR Schwyz OR Schaffhausen OR Solothurn OR St Gallen OR St. Gallen OR Ticino OR Tessin OR Thurgau OR Uri OR Vaud OR Waadt OR Valais OR Wallis OR Zug OR Zurich OR Zürich)) OR (UK OR united kingdom OR Great Britain OR Wales OR Scotland OR England OR northern ireland OR welsh OR scottish) OR ((Austria* OR Österreich*) OR (Burgenland OR Carinthia OR Kärnten OR Lower Austria OR Niederösterreich OR Upper Austria OR Oberösterreich OR Salzburg OR Styria OR Steiermark OR Tyrol OR Tirol OR Vorarlberg OR Vienna OR Wien)) OR ((Netherland OR dutch) OR (Drenthe OR Flevoland OR Friesland OR Gelderland OR Groningen OR Limburg OR North Brabant OR North Holland OR Overijssel OR south holland OR Holland OR Utrecht OR Zeeland OR Amsterdam)) OR ((Germany OR German OR Deutschland OR deutsch) OR (Baden-Wuerttemberg OR Bavaria OR Berlin OR Brandenburg OR Bremen OR Hamburg OR Hesse OR Lower Saxony OR Mecklenburg-Western Pomerania OR North Rhine-Westphalia OR Saarland OR Saxony OR Saxony-Anhalt OR Schleswig-Holstein OR Thuringia OR Thüringen OR Sachsen-Anhalt OR Sachsen OR Nordrhein-Westfalen OR Mecklenburg Vorpommern OR Niedersachsen OR Hessen OR Bayern OR Baden-Württemberg)))<br><br>AND (2011[Date - Publication] : 3000[Date - Publication]) |

Table S2. Search strategy for the database LIVIVO

| |
|---|
| MESH=(Public Health Administration OR Public Health Practice OR Personnel Turnover OR Public Health Systems Research OR Schools, Public Health OR Health Workforce OR Capacity Building OR Personnel Management OR Education, Public Health Professional)<br><br>AND TI=(public health OR health authori* OR health department* OR epidemiolog* OR public health specialt*) AND TI=(train* OR educat* OR postgrad* OR post-grad* OR specialt* OR (capacity AND building) OR specialty OR workforce* OR manpower OR competenc* OR abilit* OR skill* OR curricul* OR learning goals OR residenc*) AND TI=((((Switzerland* OR Swiss* OR Schweiz*) OR (Aargau OR Appenzell Outer-Rhodes OR Appenzell Ausser-Rhoden OR Appenzell Inner-Rhodes OR Appenzell Inner-Rhoden OR Basel OR Basel District OR Basel Landschaft OR Bern OR Bern OR Fribourg OR Freiburg OR Geneva OR Genf OR Glarus OR Glarus OR Grisons OR Graubünden OR Jura OR Jura OR Lucerne OR Luzern OR Neuchâtel OR Neuenburg OR Nidwalden OR Obwalden OR Schwyz OR Schaffhausen OR Solothurn OR St Gallen OR St. Gallen OR Ticino OR Tessin OR Thurgau OR Uri OR Vaud OR Waadt OR Valais OR Wallis OR Zug OR Zurich OR Zürich)) OR (UK OR united kingdom OR Great Britain OR Wales OR Scotland OR England OR northern ireland OR welsh OR scottish) OR ((Austria* OR Österreich*) OR (Burgenland OR Carinthia OR Kärnten OR Lower Austria OR Niederösterreich OR Upper Austria OR Oberösterreich OR Salzburg OR Styria OR Steiermark OR Tyrol OR Tirol OR Vorarlberg OR Vienna OR Wien)) OR ((Netherland OR dutch) OR (Drenthe OR Flevoland OR Friesland OR Gelderland OR Groningen OR Limburg OR North Brabant OR North Holland OR Overijssel OR south holland OR Holland OR Utrecht OR Zeeland OR Amsterdam)) OR ((Germany OR German OR Deutschland OR deutsch) OR (Baden-Wuerttemberg OR Bavaria OR Berlin OR Brandenburg OR Bremen OR Hamburg OR Hesse OR Lower Saxony OR Mecklenburg-Western Pomerania OR North Rhine-Westphalia OR Saarland OR Saxony OR Saxony-Anhalt OR Schleswig-Holstein OR Thuringia OR Thüringen OR Sachsen-Anhalt OR Sachsen OR Nordrhein-Westfalen OR Mecklenburg Vorpommern OR Niedersachsen OR Hessen OR Bayern OR Baden-Württemberg))))<br><br>AND PY=2011:2050 |

*Table S3. Overarching coding tree structure*

1. Context-related aspects of the qualification model
    - 1.1 Duration and density
    - 1.2 Professional credentialing
    - 1.3 Program objectives
    - 1.4 Recruitment and selection process
    - 1.5 Stakeholder involvement
    - 1.6 Resources
    - 1.7 Setting

2. Content-related aspects of a qualification model
    - 2.1 Integration of content into program
        - 2.1.1 Horizontal Integration
        - 2.1.2 Vertical Integration
        - 2.2 Training & education forms
        - 2.2.1 Teacher-centered training forms
        - 2.2.2 Interaction of participants
        - 2.2.3 Exposure to practice
    - 2.3 Didactical concepts
    - 2.4 Measurability and assessment

3. Translation into a qualification model
    - 3.1 Workshops, seminars and trainings
    - 3.2 Hospitation (e.g. internship) in PH-institutions
    - 3.3 Master programs in public health
    - 3.4 PhD-programs in public health
    - 3.5 Integration of PH into medical curricula
    - 3.6 Postgraduate/Trainee programs
    - 3.7 Rotation model practice and science
    - 3.8 Peer Mentoring
    - 3.9 Labelling

4. Consolidation and further development
    - 4.1 Piloting
    - 4.2 Dissemination
    - 4.3 Evaluation and quality assessment
        - 4.3.1 Evaluation methods
        - 4.3.2 Factors and barriers for program success
        - 4.3.3 Reaction (Level 1)
        - 4.3.4 Learning (Level 2)
        - 4.3.5 Behavior (Level 3)
        - 4.3.6 Results (Level 4)
        - 4.3.7 ROI (Level 5)
    - 4.4 Transferability
    - 4.5 Continuous program development

**Table S4.** Sampling plan

| Current activity in the PHS | Current activity at a university | Early career professionals | International perspective |
|---|---|---|---|
| *Federal level:* ID01, ID10, ID15, ID17 | ID02, ID03, ID04, ID08, ID09, ID13, ID14, ID16, ID20, ID21, ID23 | ID10, ID11, ID14, ID15, ID17 | ID03, ID06, ID07, ID09, ID19, ID20, ID21, ID24 |
| *State level:* ID04, ID12 | | | |
| *local level:* ID05, ID11, ID18, ID22, ID24 | | | |

Table S5. Characteristics of the studies included in the scoping review.

| First Author (Year) | Ref | Short title | Country | Qualification program, main focus |
|---|---|---|---|---|
| Baxter (2016) | (62) | A new infection trainee education programme from the Healthcare Infection Society | UK | Structured education program focusing on infection training and including infection prevention and control (IPC) |
| Bennetts (2012) | (73) | Continuing professional development for public health: an andragogical approach | UK | Continuing professional development (CPD) |
| Buunaaisie (2018) | (71) | Employability and career experiences of international graduates of MSc Public Health: a mixed methods study | UK | Master of Science Program (MSc) |
| Chastonay 2012) | (72) | Design, implementation and evaluation of a community health training program in an integrated problem-based medical curriculum | CH | 6-year longitudinal and multidisciplinary Community Health Program (CHP) |
| Cheetham (2018) | (51) | Embedded research: a promising way to create evidence-informed impact in public health? | UK | Embedded research (ER) co-located between academia and local health authority (LHA) |
| Currie (2020) | (52) | Public Health Ausbildung in Österreich. Ein Überblick | UK, AU | Specialty public health training |
| Dey (2019) | (63) | The United Kingdom Field Epidemiology Training Programme: meeting programme objectives | UK | Field epidemiology training programmes (FETP) |
| Diem (2016) | (64) | Prevention and control of noncommunicable diseases through evidence-based public health: implementing the NCD 2020 action plan | AT | Capacity building efforts focusing on knowledge translation for non-communicable diseases (NCD) |
| Dorner (2014) | (53) | Long-Term Evaluation of a Course on Evidence-Based Public Health in the U.S. and Europe | AT | Postgraduate public health university courses and public health doctoral programs |
| Erwin (2014) | (54) | Long-Term Evaluation of a Course on Evidence-Based Public Health in the U.S. and Europe | US | Evidence-based public health course |
| Gerhardus (2017) | (74) | Public Health als anwendungsorientiertes Fach und Multidisziplin – „Forschendes Lernen“ als Antwort auf die Herausforderungen für Lehren und Lernen? | DE | Research-based learning approaches within a student-led research project |
| Gillam (2016) | (69) | Public health education in UK medical schools-towards consensus | UK | Consensus statement to support the development of learning environments for public health; public health rotation |
| Gray ( 2018) | (55) | Developing the public health workforce: training and recognizing specialists in public health from backgrounds other than medicine: experience in the UK | UK | Multidisciplinary training of senior public health specialists;<br><br>Registry for public health specialists |
| Harrison (2015) | (56) | The effect of using different competence frameworks to audit the content of a masters program in public health | UK | Competence frameworks in master public health programs |
| Heller (2017) | (57) | Open Online Courses in Public Health: experience from Peoples-uni | UK | Open Online Courses (OOCs) as open educational resources |
| Idler (2016) | (58) | Prevention and health promotion from theory to practice: The interprofessional MeMPE Summer University for students of Medicine, Master of Public Health and Epidemiology | DE | Interprofessional Seminar on prevention and health promotion in the course of a summer university |

| | | | | |
|---|---|---|---|---|
| Jansen (2013) | (59) | A masterclass to teach public health professionals to conduct practice-based research to promote evidence-based practice: a case study from The Netherlands | NL | Masterclass for public health professionals focusing on practice-based research skills |
| Könings (2018) | (60) | Is blended learning and problem-based learning course design suited to develop future public health leaders? | NL | Problem-based, blended learning method in a virtual learning environment |
| McCulloch (2014) | (67) | Developing capacity in field epidemiology in England | UK | Field epidemiology training programmes (FETP) |
| Peik (2016) | (65) | Comparison of public health and preventive medicine physician specialty training in six countries: Identifying challenges and opportunities | US | Descriptive profiles of national training demographics and structures of Public health and preventive medicine (PHPM) specialties |
| Ramsay (2014) | (61) | The Structured Operational Research and Training Initiative for public health programmes | CH | Structured operational research and training initiative |
| Salway (2013) | (76) | Improving capacity in ethnicity and health research: report of a tailored programme for NHS Public Health practitioners | UK | Tailored program focusing on research capacity development |
| Smith (2015) | (68) | Principles of all-inclusive public health: developing a public health leadership curriculum | UK | Curriculum development focusing on public health leadership skills |
| Tran (2017) | (75) | In-house peer supported literature search training: a public health perspective | UK | Peer supported literature search training course; Knowledge and library service (KLS) |
| Turner-Wilson (2017) | (70) | Can nurses rise to the public health challenge? How a novel solution in nurse education can address this contemporary question | UK | Public health improvement theme running throughout an undergraduate nursing curriculum |

2
